# Global patterns and correlates in the emergence of antimicrobial resistance in humans

**DOI:** 10.1101/2022.09.29.22280519

**Authors:** Emma Mendelsohn, Noam Ross, Carlos Zambrana-Torrelio, T. P. Van Boeckel, Ramanan Laxminarayan, Peter Daszak

## Abstract

Antimicrobial resistance (AMR) is a critical global health threat, and drivers of the emergence of novel strains of antibiotic-resistant bacteria in humans are poorly understood at the global scale. We examined correlates of AMR emergence in humans using global data on the origins of novel strains of AMR bacteria from 2006 to 2017, human and livestock antibiotic use, country economic activity, and reporting bias indicators. We found that AMR emergence is positively correlated with antibiotic consumption in humans, whereas the relationship with antibiotic consumption in livestock is modified by gross domestic product (GDP), with only higher GDP countries showing a slight positive association. We also found that human travel may play a role in AMR emergence, likely driving the spread of novel AMR strains into countries where they are subsequently detected for the first time. Finally, we produced predictive models and country-level maps of the global distribution of AMR risk. We assessed these against spatial patterns of reported AMR emergence, to identify gaps in surveillance that can be used to direct prevention and intervention policies.

## Introduction

The emergence of antimicrobial resistance (AMR) is a critical global health challenge. AMR bacterial strains have been associated with increased mortality, longer illnesses, medical complications in surgery, barriers to chemotherapy, and higher health care costs (Cosgrove 2006; World Health Organization 2014, 2015; Interagency Coordination Group on Antimicrobial Resistance 2019). Global human use of antibiotics has increased substantially over the last two decades, with an alarming uptick in last-resort compounds that are administered when other treatments fail (Klein et al. 2020). Rates of human use of antibiotics correlate with resistance rates in pathogenic bacteria at multiple scales and locations (Goossens et al. 2005; Riedel et al. 2007; Bell et al. 2014; Llor and Bjerrum 2014). Combating AMR has become a priority for governments (e.g., the United States National Action Plan for Combating Antibiotic-Resistant Bacteria; UK Five-Year National Action Plan for Tackling Antimicrobial Resistance; Australia’s National Antimicrobial Resistance Strategy) and intergovernmental organizations (e.g., Tripartite-Plus Alliance on AMR, Food and Agriculture Organization, World Organisation for Animal Health, United Nations Environment Programme, and World Health Organization) and multi-lateral development banks and financing facilities (e.g., World Bank).

Antibiotics are used routinely in livestock to prevent and treat bacterial diseases, and as growth promoters to expedite weight gain (Newell et al. 2010; Van Boeckel et al. 2019). Antibiotic use in livestock—which vastly exceeds their use in humans—has enabled intensive husbandry, and is projected to increase by 67% globally, and to nearly double in Brazil, Russia, India, China, and South Africa by 2030 (Van Boeckel et al. 2015; Van Boeckel et al. 2019). Resistance genes, AMR bacterial strains, and plasmids including some of human clinical relevance, such as MRSA (Methicillin-resistant *Staphylococcus aureus*), have been reported from livestock, wildlife, and environmental samples (Van Boeckel et al. 2019; Wellington et al. 2013; Papadopoulos et al. 2018; European Food Safety et al. 2019; Tsai et al. 2020). These findings have led to policy efforts to reduce antibiotic use in livestock (World Health Organization 2015; Interagency Coordination Group on Antimicrobial Resistance 2019). However, the relative roles of antibiotic use in animals or humans in driving AMR emergence of clinical relevance to humans, has not yet been thoroughly assessed.

To our knowledge, there are no published analyses on the relative roles of human and livestock consumption of antibiotics in driving the emergence of novel strains of antibiotic-resistant bacteria in human clinical cases. In the current study, we use a database that we assembled of global AMR emergence events containing 1,604 records of the first clinical reports of novel bacterial resistance over 11 years from 2006 to 2017, to examine global patterns in the emergence of new AMR strains in humans (Mendelsohn et al. 2021). We model how observed AMR emergence events are correlated with human and livestock consumption of antibiotics; human population and mobility (migrant population and tourism); economic activity (gross domestic product [GDP] and healthcare expenditure); antibiotic exports as a proxy for production; and biomedical surveillance efforts. We then use these correlations to produce predictive models of the global distribution of AMR emergence risk.

A key challenge in interpreting global patterns of AMR emergence is variation in AMR surveillance and reporting. Underreporting in lower-income countries is a persistent problem in AMR datasets (World Health Organization 2014; ResistanceMap 2017), and may be particularly important given that many lower-income countries are most affected by resistant infections (e.g., malaria, tuberculosis, neonatal sepsis) (Byarugaba 2004; World Health Organization 2015; Laxminarayan et al. 2016) and are experiencing the greatest increases in consumption of antibiotics in humans and livestock (Klein et al. 2018; Van Boeckel et al. 2019). In this study, we apply methods used in our previous work analyzing the emergence of zoonoses (Allen et al. 2017) to correct for underlying biases in reporting novel emergence by using quantitative metrics of AMR surveillance.

## Results

Our AMR emergence database contains 1,604 records of first clinical reports of novel bacterial resistance occurring in 59 countries from 2006 to 2017, extracted from biomedical literature. The United States had the greatest number of reported events (n = 132), followed by India (n = 127), China (n = 120), Canada (n = 98) and Japan (n = 75). For more detail on the database, see Mendelsohn et al. 2021.

We modeled the frequency of reported AMR emergence as a function of human antibiotic consumption, animal antibiotic consumption, per capita GDP, health care expenditure (% of GDP), population, inbound tourism and migrant population, and measures of reporting and publication bias. Our model explained 63% (standard deviation = 5.1%) of country-level variance in AMR emergence rates.

Human antibiotic consumption was positively associated with AMR emergence rates (odds ratio [OR] = 1.04 per defined daily dose [DDD]; 89% credible interval [89CI] = 1.00-1.10) (**Figure 1**). We use 89% as the range for credible intervals because it is considered more stable than higher ranges, such as the commonly used 95% intervals (Makowski et al. 2019). For a country in which AMR emergence is expected (i.e., non-zero prediction), a 33% (±6%) greater than average AMR emergence rate is expected at twice the average human antibiotic consumption (mean human antibiotic consumption = 7.5 DDD), with all other variables held at average.

**Figure 1.**
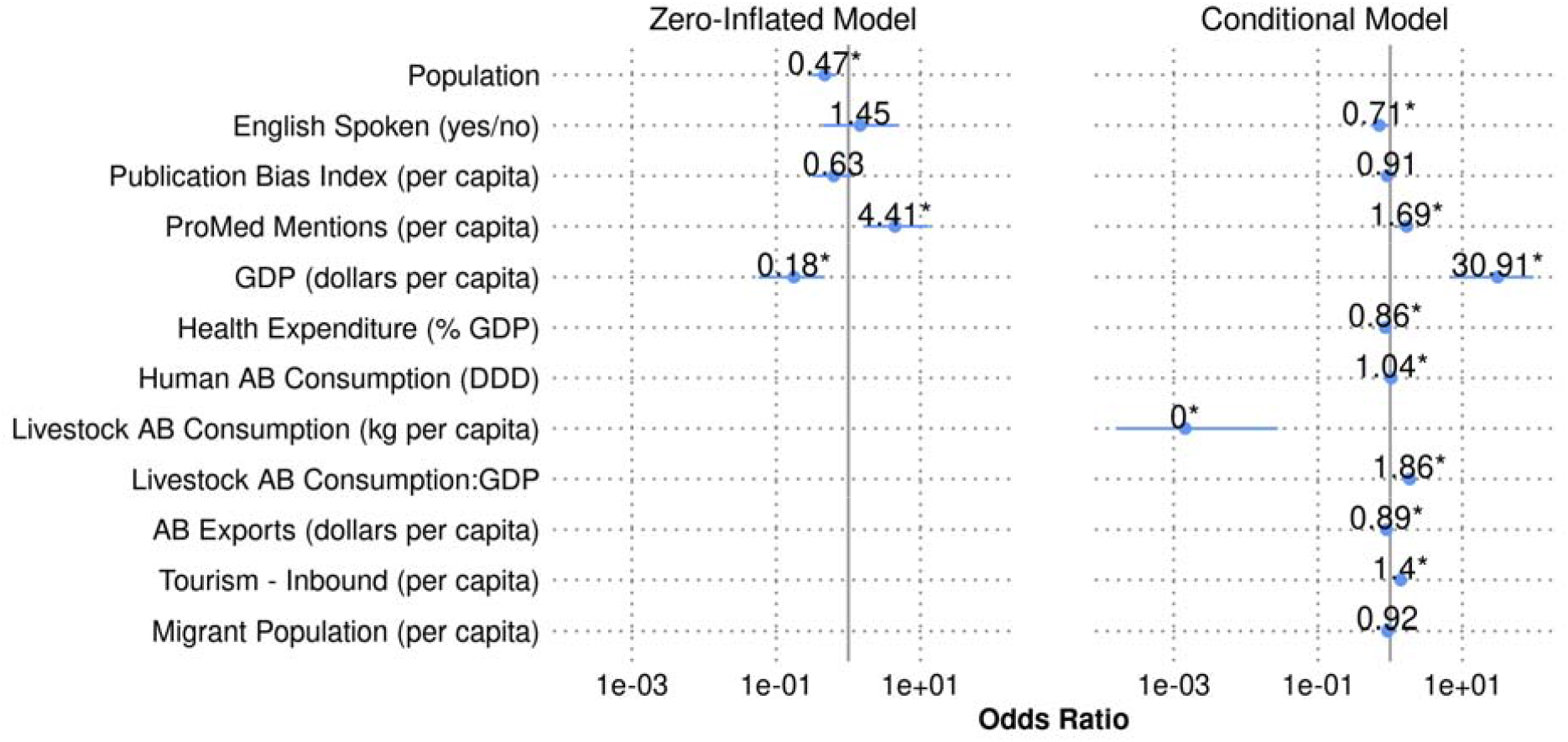
Odds ratios and 89% credible intervals of features in the main model. Asterisks indicate that the variable is a consistent predictor of the outcome (i.e., 89% credible intervals do not include 1).

The interaction between livestock antibiotic consumption and GDP, both normalized to the human population, was a consistent predictor of AMR emergence rates (OR = 1.9; 89CI = 1.4-2.3) (**Figure 1**). This interaction term indicates that the effect of livestock antibiotic consumption on AMR emergence increases at increasing levels of GDP (**Figure 2**). Specifically, for every unit increase in GDP (log-dollars per human capita), the log-odds of the effect of livestock antibiotic consumption (logged, kg per human capita) increases by log(1.9). The main (non-interaction) effect of livestock antibiotic consumption was consistently inversely associated with AMR emergence (OR = 0.0014 per log of kg antibiotics consumed by livestock per human capita; 89CI = 0.00016-0.028), which accounts for the negative relationship between livestock antibiotic consumption and AMR emergence observed at lower GDP levels. The overall effect of GDP (log-dollars per human capita) was highly associated with AMR emergence (OR = 31; 89CI = 7.2-97).

**Figure 2.**
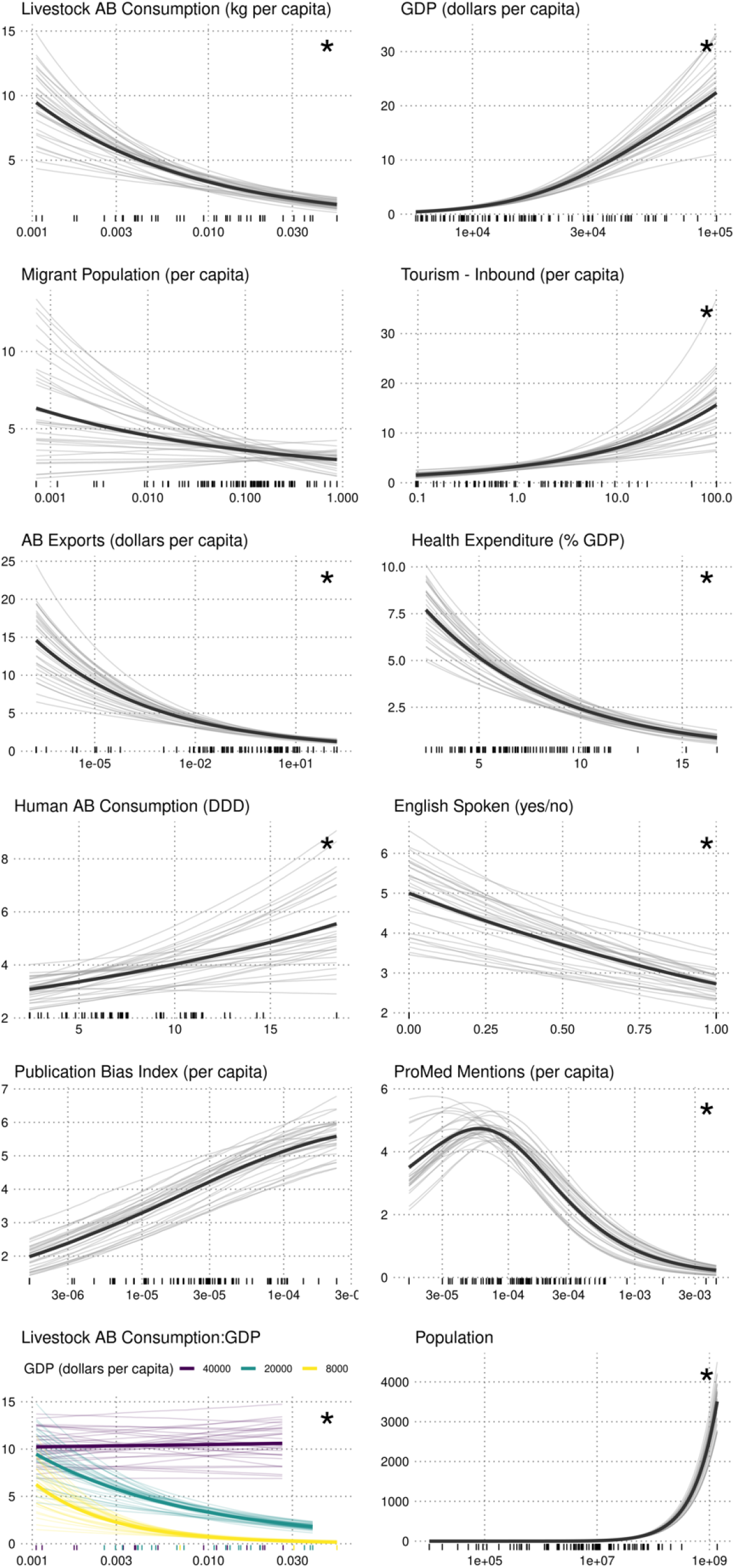
Additive change in counts (marginal effects) of AMR emergence for each model variable. Faded lines represent individual model iterations; solid line is average model. Rug ticks show raw values. Asterisks indicate that the variable is a consistent predictor of the outcome (i.e., 89% credible intervals of odds ratios do not include 1).

Inbound tourism volume per country, normalized to population, was positively associated with AMR emergence (OR = 1.4 per log of inbound tourists per capita; 89CI = 1.2-1.6). For a country in which AMR emergence is expected (i.e., non-zero prediction), a 26% (±5%) greater than average AMR emergence rate is expected at twice the average inbound tourism (mean inbound tourism per capita = 1.4), with all other variables held at average. Migrant population, normalized to total population, was not a consistent predictor of the outcome (OR = 0.92 per log of migrant population per capita; 89CI = 0.76-1.1). The dollar value of antibiotic exports, normalized to the human population, was consistently inversely associated with AMR emergence (OR = 0.89 per log of antibiotic exports per capita; 89CI = 0.87-0.93). Healthcare expenditure was also consistently inversely associated with AMR emergence (OR = 0.85 per percent of GDP; 89CI = 0.83-0.90).

We used several variables to quantify reporting bias in AMR reports: The number of times a report of an AMR disease on ProMED related to a country (ProMED mentions) was consistently positively associated with AMR emergence (OR = 1.7 per log of ProMED mentions per capita; 89CI = 1.2-2.1), while speaking English in a country was inversely related (OR = 0.71; 89CI = 0.53-0.96), and the publication bias index was not a consistent predictor (OR = 0.91, 89CI = 0.81-1.0). Partial effect plots in both parts of the hurdle model are shown in **Figure S1**.

Due to lack of data on human and livestock antibiotic consumption for many, especially low-income countries, (**Table S1**), we used model-imputed values for these correlates. To test robustness of results, we evaluated results under four imputation scenarios: 1) no imputation of antibiotic consumption (n = 36), 2) imputation of either human or livestock antibiotic consumption (n = 73), 3) imputation of human and livestock antibiotic consumption for countries within GDP range of countries with complete human and animal antimicrobial consumption data (n = 88), and 4) full imputation (n = 190). While odds ratios differed among the models, the overall direction of effects was consistent, and interpretation did not vary drastically between models (**Figure S2**). For reporting model results here, we use the third scenario—imputation of human and livestock antibiotic consumption for countries within the GDP range of countries with complete human and animal antimicrobial consumption data. This scenario was selected because it maximizes data coverage without predicting beyond the conditions of the observed data. Results for all other scenarios are reported in the Supplementary Information.

We also tested alternative formulations of the model to assess the robustness of results. Because the United States is a singular outlier in the number of reported events, GDP, publication bias index, ProMED mentions, and antibiotic exports, we ran a model without the United States. In this scenario, use of the English language in a country and healthcare expenditure were no longer associated with AMR emergence, the publication bias index became inversely associated, and other results remained largely the same (**Figure S2**). In a separate scenario, we replaced per-human-capita livestock antibiotic consumption with per-livestock biomass antibiotic consumption and found that per-livestock biomass antibiotic consumption was not associated with AMR emergence while livestock population on its own was inversely associated with emergence. Finally, we repeated the analysis on a subset of emergence data representing the first global appearances of unique drug-pathogen combinations (i.e., including only the first country in which resistance of a pathogen to a drug is observed). Results were largely consistent with the main model, with the use of the English speaking in a country becoming no longer associated with AMR emergence and the publication bias index becoming positively associated.

We used our model to estimate zero-corrected AMR emergence rates for each country, that is, predicted rates conditional on equal reporting variables across countries (**Figure 3**). These results show higher predicted rates for 77% of countries, including those that have the highest counts in our database (United States, China) and in countries that previously reported few or zero events. Countries with the greatest increase in predicted AMR counts were Russia (95^th^ percentile range = 109-367), Saudi Arabia (91-379), and Turkmenistan (19-96), all of which had zero reported events in our database.

**Figure 3.**
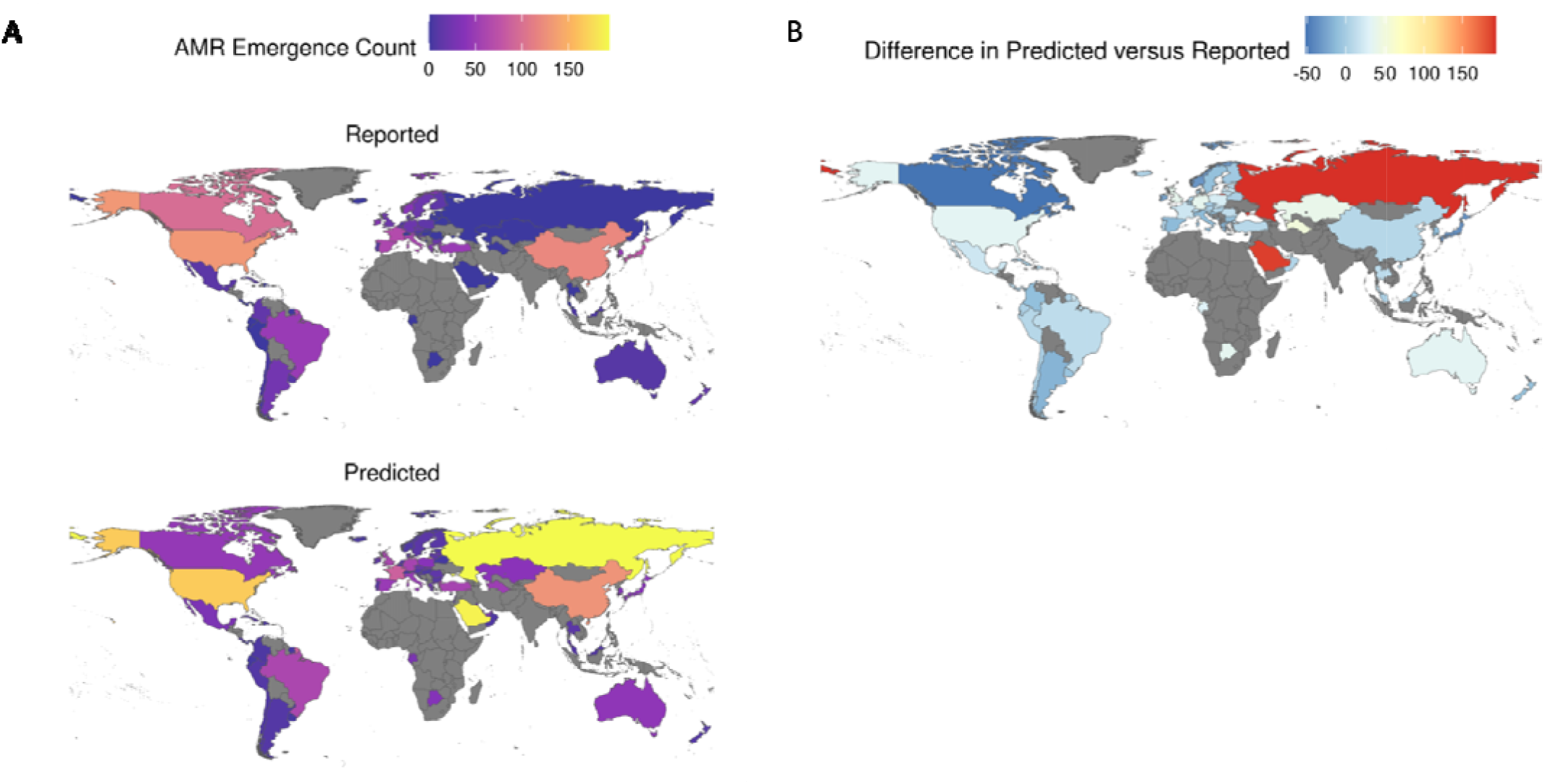
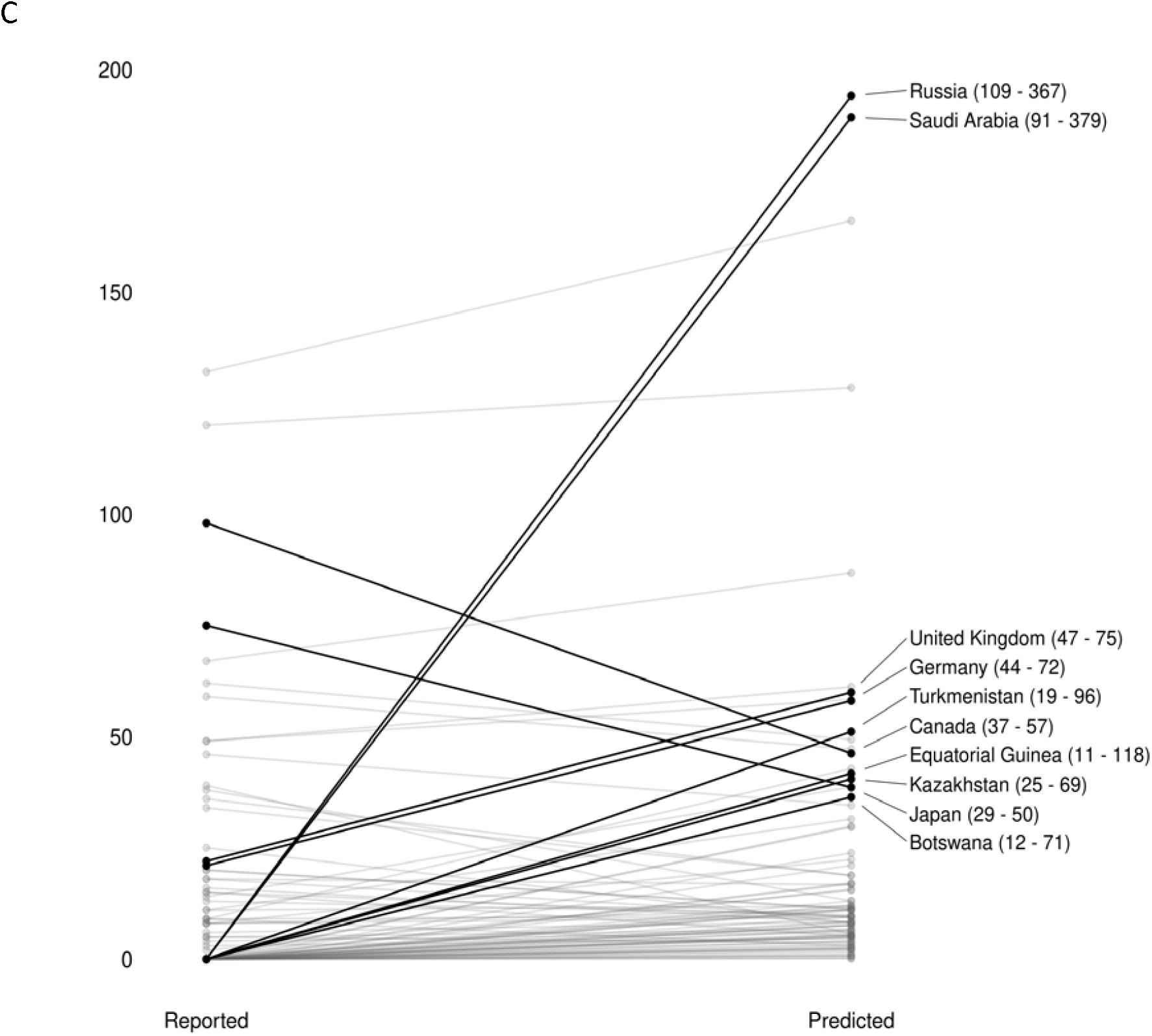
Reported and median predicted AMR emergence event counts for 88 countries (A). Difference between predicted and reported counts (B). 10 countries with largest absolute difference between reported and median predicted counts, with 95^th^ percentile predicted range in parenthesis (C).

## Discussion

This paper reports the first global analysis of drivers of the emergence of antimicrobial resistance (AMR) in humans, with efforts to correct for reporting bias and inconsistencies in data on antibiotic use. Previous studies have described the presence, prevalence of, and trends in caseloads over time for specific resistant strains (Riedel et al. 2007; Song et al. 2011; Smith et al. 2013; Llor and Bjerrum 2014; Paterson et al. 2014; Papadopoulos et al. 2018). Others have reviewed broad patterns in the emergence of AMR based on trends in the literature without correcting for underlying ascertainment bias, or testing hypotheses on underlying causal factors (Byarugaba 2004; Bonn 2007; Bell et al. 2014; World Health Organization 2014). Some studies have analyzed patterns of use or sale of antibiotics for human or livestock use in specific regions or globally (Goossens et al. 2005; Van Boeckel et al. 2015; Klein et al. 2018). There is a previous analysis of broad patterns of global AMR emergence in livestock that evaluated surveillance bias but did not test hypotheses on the relative significance of different drivers (Van Boeckel et al. 2019). Efforts to identify trends and drivers of emerging infectious diseases are hampered by a lack of clarity on the origins of past events, and by spatial and temporal biases in surveillance (Jones et al. 2008; Allen et al. 2017). Here, we used records of first clinical reports of unique bacterial-drug cases from 2006 to 2017 (Mendelsohn et al. 2021), datasets of antimicrobial drug sales for human and livestock use, and published strategies for dealing with reporting bias, to analyze the origins, trends, and likely drivers of global emergence of AMR.

Our analysis showed that human use of antimicrobials is positively correlated with the origins of AMR events in people, and that this scales with defined daily dose (DDD). Previous analyses of AMR trends have modeled the presence or prevalence of specific resistant strains and provided evidence that antibiotic use in people directly contributes to AMR in hospitals and clinics, communities, and countries (Goossens et al. 2005; Riedel et al. 2007; Koningstein et al. 2010; Bell et al. 2014; Llor and Bjerrum 2014). However, this correlation has not previously been demonstrated on a global scale, controlling for reporting biases, and over a broad swath of AMR pathogen/drug combinations. Another prior study assessed how socioeconomic and demographic factors correlate with an index of antimicrobial resistance in 103 countries and found that human antimicrobial drug use was not correlated with resistance (Collignon et al. 2018). However, the current study analyzes the drivers of the first known clinical cases of a novel AMR emergence, whereas (Collignon et al. 2018) analyzed the level of resistance to several drug classes in three pathogens encountered in clinics in a country. Our analysis is consistent with the findings of (Collignon et al. 2018) that the prevalence of AMR in a country is likely driven also by contagion – the spread of antimicrobial resistance after its emergence – and that this occurs independently to the degree of antibiotic consumption. Together, these papers provide a more detailed explanation of what drives the origins, spread and impact of AMR, and are therefore of value in developing policy to control each aspect of emergence.

We found that the relationship between antibiotic consumption for animal husbandry and the origins of new AMR strains in people is modified by GDP, with the highest GDP countries having a slight positive association, and lower GDP countries having a neutral or negative association. Under a separate formulation, in which we normalized livestock antibiotic consumption to livestock biomass instead of human population, no consistent association with AMR emergence was observed. Given the limited number of data points for livestock antibiotic consumption (n = 41), additional data collection is needed to better understand the relationship between animal husbandry and AMR emergence. Nonetheless, our findings suggest that the relationship between antibiotic use in livestock and the emergence of novel AMR strains in humans may be complex or mediated by other factors.

Other research has demonstrated that antibiotic use for animal husbandry is a significant public health threat in contributing to the spread of specific existing resistance strains in humans (Vieira et al. 2011; Smith et al. 2013). A meta-analysis of antimicrobial use in animals that includes a small number (n=21) of human AMR cases, found that reduced animal use of antimicrobials led to a reduction in the pooled prevalence of AMR cases in people (Tang et al. 2017). We conclude that, while our analysis indicates that human use of antibiotics is likely more important for human AMR emergence than animal use, further research is needed to better understand the patterns of transmission of AMR strains among livestock and people (Vieira et al. 2011; Smith et al. 2013; Muloi et al. 2018). We hypothesize that dense populations of livestock may act as maintenance or amplifying hosts for known AMR strains, a scenario similar to the role of intermediate livestock hosts in the emergence of novel zoonoses such as Nipah virus disease, MERS and SARS (Morse et al. 2012).

In recent years, environmental contamination by antibiotics has been increasingly linked to the emergence and spread of AMR (Wellington et al. 2013; Singer et al. 2016). In our analysis, we assumed that countries with higher levels of production (and therefore export) of antibiotics would have higher environmental contamination. Countries with antibiotic export had lower rates of AMR emergence, suggesting that either this is a poor proxy, or that environmental contamination is not a significant driver of novel strain emergence. This does exclude the possibility of environmental contamination being a factor in maintaining or spreading AMR strains once they have emerged.

To assess if the emergence of a novel strain is caused by the spread of infection (bacterium or gene transmission) into a country rather than its *de facto* evolution and origin, we included measures of human population movement in our model. Inbound tourism, normalized to population, was a predictor of AMR emergence, while inbound migration, normalized to population, was not. These results suggest that first emergences in a country may be driven, in part, by the spread of existing resistant strains from other countries. We repeated the analysis on a subset of emergence data representing first global appearances of unique drug-pathogen combinations (i.e., including only the first country in which resistance of a pathogen to a drug is observed). This analysis did not alter our findings related to tourism and migration, suggesting that mechanisms of spread (e.g., gene transfer) in addition to mutation may also drive first global emergences.

More developed public infrastructure and higher metrics of good governance inversely correlate with AMR rates (Collignon et al. 2018). In our study, we used data on GDP per capita and healthcare expenditure as proxies for the ability of countries to control AMR, identify cases, and manage consumption patterns through education programs. GDP per capita was consistently positively associated with AMR emergence in a country. Healthcare expenditure was consistently inversely associated with AMR emergence in a country, but this relationship was no longer consistent when we removed the United States from the dataset. These findings likely reflect the fact that our outcome measure is not the level of resistance seen in clinics in a country (e.g. prevalence, incidence, occurrence of known or novel AMR strains), but the number of novel AMR strains originating in a country. The latter may be more strongly correlated with human antibiotic drug use as a driver of the evolution and emergence of novel strains, while the former is linked to ability to control these strains once they have emerged.

In previous work, we analyzed global trends and identified predictive hotspots of emerging infectious diseases (Jones et al. 2008), and emerging zoonoses (Allen et al. 2017) by correcting for underlying biases in reporting of novel emergence. In the current study, we accounted for country-level surveillance and reporting effort by including use of English language in a country (as the database was limited to English-language literature), number of ProMED mentions, and a publication bias index produced previously (Allen et al. 2017). We used our model to estimate predicted (zero-corrected) AMR rates of emergence for each country, and found the greatest increase in predicted rates in Russia, Saudi Arabia, and Turkmenistan, all of which had zero reported events. These findings point to significant reporting gaps in these countries and the need to apply surveillance beyond the relatively limited number of countries where surveillance currently occurs.

There are several limitations to this study. First, it analyzes trends in novel AMR strains reported in the literature from 2006 to 2017 against data on potential drivers from different time periods within this range. Variation in these factors over the 11 years of AMR reporting may reduce the accuracy of the analysis. This may be further confounded if countries that identify novel AMR events have then significantly reduced or modified antimicrobial use. Second, it uses published data on novel AMR strains. While we included several measures of reporting and publication bias, the changes in interest or capacity to diagnose and identify AMR over this period may have varied among countries irrespective of economic capacity, due to trends in research fields. Third, data availability of some of the correlates are skewed to richer countries. Livestock antibiotic consumption data, estimated from country-reported antibiotic sales from livestock (Van Boeckel et al. 2019), is especially sparse (available for 41 countries) and biased towards developed economies. Finally, it is important to emphasize that the relationships discussed in this paper are associative, and causality can only be hypothesized through this type of global analysis. Further work on the mechanisms of what drives the origin of new strains, and what drives their maintenance, amplification and spread is urgently needed.

## Methods

### Data

We used AMR emergence data from the database described in (Mendelsohn et al. (2021), (https://zenodo.org/record/4924992) which contains records of first clinical reports of unique bacterial-drug AMR detections from 1998-2017, drawn from scientific literature and disease surveillance reports. We filtered the database for events starting in 2006 and later, as database coverage prior to 2006 is limited to disease surveillance reports. To perform analyses at the country level, we summed the count of emergence events by country. This approach allows the same drug-bacteria combination to be represented in multiple country counts. As part of our robustness analysis (below), we also ran the model using first reported global emergences as an alternate outcome (i.e., each drug-bacteria combination reported only once).

Predictor variables are from multiple sources, listed in **Table S1**. We included data on human and livestock consumption of antibiotics, human population and mobility (migrant population and tourism), economic activity (GDP and healthcare expenditure), and antibiotic exports as a proxy for production. In addition, we included five variables representing reporting bias: population, GDP, English language spoken, ProMED mentions, and publication bias index. The publication bias index is based on total biomedical publications originating from or referring to geographic regions, (Allen et al. 2017), an approach used for a variety of global-scale disease detection studies (Huff et al. 2016; Olival et al. 2017; Carlson et al. 2022).

Prior to modeling, lognormally distributed continuous variables were natural log transformed, and some variables were normalized to GDP or population, as indicated in the *measurement units* field of **Table S1**. Livestock antibiotic consumption was normalized to human population, rather than livestock population, as we are interested in the potential contribution of antibiotic use in agriculture to AMR emergence in humans. In our robustness analysis (below), we ran an alternate version of the model with livestock antibiotic consumption normalized to livestock biomass. Because initial data exploration found that the relationship between livestock antibiotic consumption and AMR emergence differed between low- and high-income countries, we included an interaction term for livestock antibiotic consumption and country GDP.

### Missing data handling

We limited the total number of countries in the dataset to those that have population and GDP data available (n = 190). As shown in **Table S1**, data availability was not consistent across other variables. We inferred zeros for missing values for the AMR emergence field, and one half the minimum value for the publication bias index, ProMED mentions, and antibiotic export fields.

The following remaining variables were unavailable for some countries, with a distinct bias of missing data in low-income countries: human antibiotic consumption, livestock antibiotic consumption, health expenditure, and inbound tourism. We imputed missing values for these variables, using four approaches of to check for robustness:

1. *No imputation of antibiotic consumption* – Dataset limited to countries with values for both human *and* animal antimicrobial consumption (n = 36).
2. *Imputation of either human or livestock antibiotic consumption* – Dataset includes countries with values for human *and*/*or* animal antibiotic consumption (n = 73).
3. *Imputation of human and livestock antibiotic consumption for countries within GDP range* – Dataset includes countries that are missing both human and livestock antibiotic consumption *if* the country has a GDP within the range of GDPs of countries from scenario 1 ($5,870/capita [Thailand] - $101,417/capita [Luxembourg]), which have both human *and* animal antimicrobial consumption data (n = 88).
4. *Full imputation* – Includes all countries in the dataset (n = 190).

We used a Multivariate Imputation by Chained Equations (MICE) algorithm with classification and regression trees (CART) to model missing values based on the available data (Buuren 2018). CARTs are commonly used for imputation for their robustness against outliers and ability to handle multicollinearity and skewed distributions (Buuren 2018). For each variable we generated 30 imputations, each with 40 iterations. We visually examined diagnostic plots to confirm convergence. We included two additional variables—antibiotic imports and livestock biomass—in the MICE routine to better estimate missing consumption data. We did not include these variables in the model itself, however, as consumption is a better estimate of direct antibiotic exposure.

### Robustness scenarios

We tested several alternative formulations of our model to determine robustness of results. We used scenario 3 for missing data handing (see above; n = 88) for all robustness scenarios. First, because the United States is a singular outlier in the number of reported events, GDP, publication bias index, ProMED mentions, and antibiotic sales (which informs the human and livestock antibiotic consumption variables), we ran a model with the United States removed. Second, we used an alternative scaling of livestock antibiotic consumption, replacing per-human-capita livestock antibiotic consumption with per-livestock biomass antibiotic consumption. In this formulation, we also included livestock biomass, defined as the total mass of cattle, pig, and chicken populations within a country (Van Boeckel et al. 2015), as a separate feature to allow disaggregation of the effects of livestock antibiotic consumption and livestock biomass. Finally, we used an alternate outcome variable representing the first global emergence of antibiotic strains within countries, rather than first national emergences. This way, a drug-bacteria combination is counted only in the single country in which it first emerged in our dataset.

### Statistical approach

We first examined data for collinearity, and spearman rank coefficients were less than 0.7 for all variable pairs. We used a Poisson-hurdle model, in which the response is a function of two components: a logistic component representing the probability of observing any reported cases, and a Poisson component of the number of AMR emergence events in the period, conditional on observed reporting. For the logistic equation, we included our five reporting bias variables (population, GDP, English language spoken, ProMED mentions, and publication bias index). All variables were included in the Poisson component, with population treated as an offset variable.

We took a Bayesian approach to estimating model parameters, using a No-U-Turn Hamiltonian Monte Carlo implemented in Stan and assuming a wide student’s t-distribution (nu = 3, mu = 0, sigma = 10) for all coefficient priors (Hoffman and Gelman 2014; Stan Development Team 2018). We used four Markov chains with 2000 iterations per chain to fit the model on each of the multiple imputed datasets. Posterior samples were then combined across all imputed datasets to generate posterior distributions.

We visually examined Markov chain trace plots and used effective sample convergence statistics (R-hat convergence <1.05) to confirm convergence across chains (Vehtari et al. 2019). We compared posterior predictions to the empirical distribution of AMR events with density overlay plots and interval plots. In addition, we compared the proportion of zeros in the posterior predictions to the empirical proportion to confirm that hurdle model accurately captured the excess zeros in the dataset (**Figure S3**).

### Model Predictions

We generated zero-corrected predictions of AMR emergence counts by calculating predictions for all countries our raw dataset (including imputed values) using only the Poisson component of the model, assuming a reporting probability of 1. By removing the logistic equation from the hurdle model, we were able to estimate AMR emergence counts corrected for excess zeros due to underreporting. We used a sample of 500 beta coefficients generated from our model for each variable to be able to produce median and 95^th^ percentile count estimates for each country.

### Software and reproducibility

Data analysis was performed in R version 4.0.4 (R Core Team 2021), using the tidyverse framework for data manipulation (Wickham 2017) and the drake package for workflow design (Landau 2018). We used the mice package (Groothuis-Oudshoorn 2011) for the MICE imputation routine and the brms package (Bürkner 2017), built on the Stan language (Stan Development Team 2018) for Bayesian model fitting. Visual model diagnostics were generated with the bayesplot package (Gabry and Mahr 2019).

All code and data used in this project are available for download at https://github.com/ecohealthalliance/amr-analysis and on Zenodo (https://zenodo.org/record/7051952).

## Data Availability

https://zenodo.org/record/7051952

## Author contributions

C.Z-T. and P.D. conceived the study. E.M. and N.R. developed the model and wrote code. E.M., N.R., and P.D. drafted the manuscript. R.L, and TVB provided expert review. All authors were involved in editing and approving the manuscript.

## Acknowledgments

This work was made possible by the generous support of the American people through the United States Agency for International Development (USAID) Emerging Pandemic Threats PREDICT (Cooperative Agreement No. AID-OAA0A-14-00102). The contents are the responsibility of the authors and do not necessarily reflect the views or the policy of USAID or the United States Government.

