## Supplemental Material for "Global patterns and correlates in the emergence of antimicrobial resistance in humans"

|  | **Reporting bias variable** | **Measurement Units** | **N countries with data** | **Missing data approach** | **Source** | **Year(s)** | **Note** |
| --- | --- | --- | --- | --- | --- | --- | --- |
| AMR emergence events |  | Count | 59 | Infer zero | Mendelsohn et al. 2021^1^ | 2006 - 2017 |  |
| Population | x | Count | 190 | N/A – no missing data | World Bank^2^ | 2015 | Includes all country residents regardless of legal status or citizenship |
| GDP | x | US dollars per capita | 190 | N/A – no missing data | World Bank^2^ | 2015 | Dollar figures are converted from domestic currencies using single year official exchange rates |
| English spoken in country | x | Yes/No | 190 | N/A – no missing data | CIA World Factbook^3^ | 2019 | Any country where English is widely spoken, even if not official language |
| Publication bias index | x | per capita | 157 | Infer ½ minimum value (for log transformation) | Allen et al. 2017^4^ |  | Estimated relative reporting effort in each country based on location mentions in PubMed Central Open Access Subset (PMCOAS) |
| ProMED mentions | x | per capita | 189 | Infer ½ minimum value (for log transformation) |  |  |  |
| Antibiotic exports |  | US dollars per capita | 135 | Infer ½ minimum value (for log transformation) | COMTRADE via Observatory of Economic Complexity^5^ | 2015 |  |
| Antibiotic imports* |  | US dollars per capita | 178 | N/A – variable included only to support MICE | COMTRADE via Observatory of Economic Complexity^5^ | 2015 |  |
| Health expenditure |  | % GDP | 184 | Multivariate Imputation by Chained Equations (MICE) | World Health Organization via World Bank^2^ | 2015 | Includes all healthcare goods and services consumed during each year |
| Migrant population |  | per capita | 190 | N/A – no missing data | United Nations Population Division via World Bank^2^ | 2015 | Number of people born in a country other than that in which they live, including refugees |
| Tourism (inbound) |  | per capita | 111 | Multivariate Imputation by Chained Equations (MICE) | World Tourism Organization^6^ | 2015 | Data compiled from administrative records and/or border surveys |
| Human antibiotic consumption |  | Defined Daily Dose (DDD) | 68 | Multivariate Imputation by Chained Equations (MICE) | ResistanceMap^7^ | 2014 | Data compiled from industry sources |
| Livestock antibiotic consumption |  | Kg per capita | 41 | Multivariate Imputation by Chained Equations (MICE) | Van Boeckel et al. 2019^8^ | 2010 | Country-reported antibiotic sales for cattle, chickens, pigs |
| Livestock population* |  | Population Correction Unit (PCU) | 176 | N/A – variable included only to support MICE | Van Boeckel et al. 2019^8^ | 2010 | Represents biomass of domestic cattle, chickens, pigs |

**Table S1.** Model data sources

* Variable not included in hurdle model but used to model missing data with Multivariate Imputation by Chained Equations (MICE) algorithm

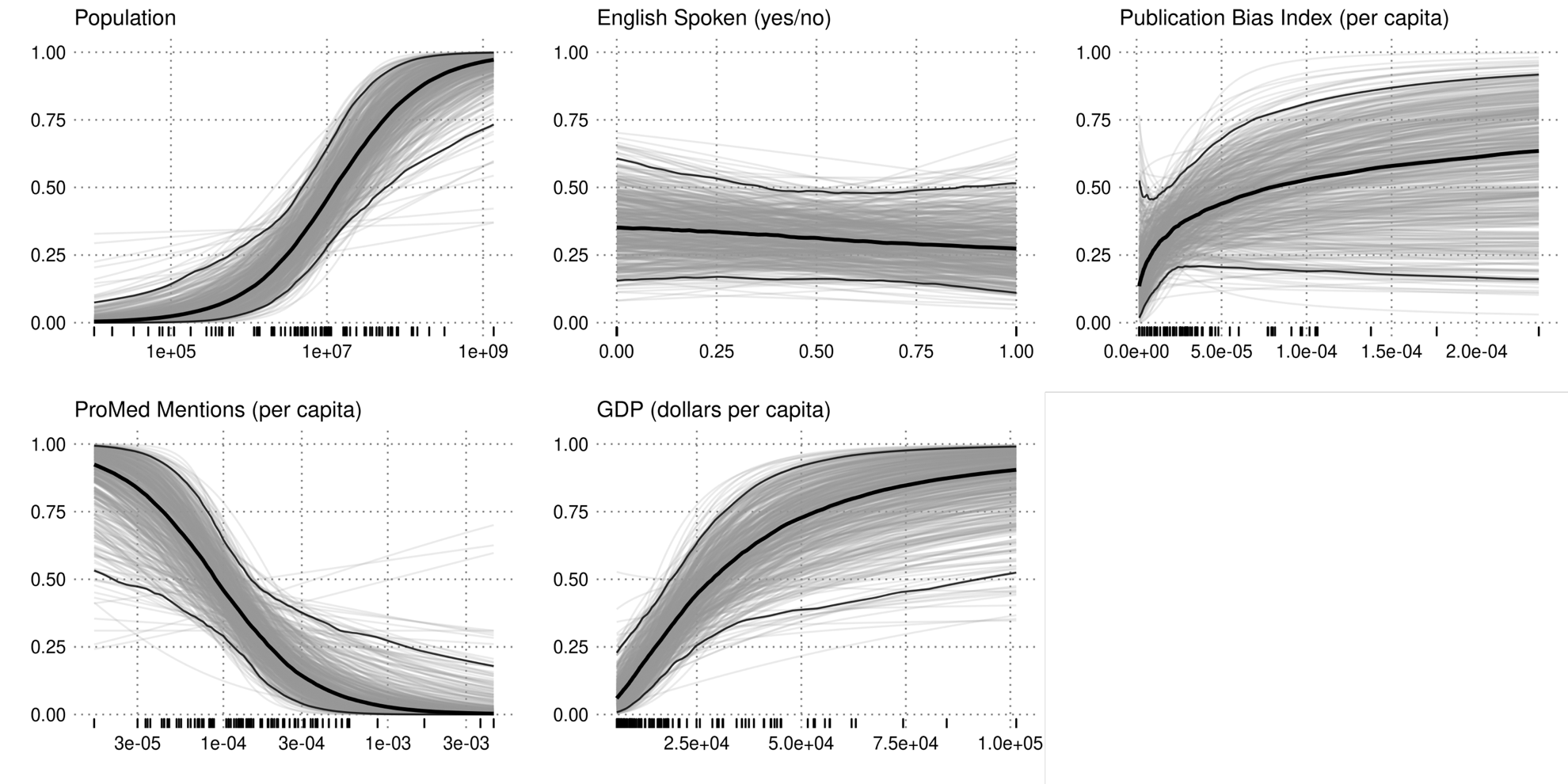

**Figure S1-a.** Partial effect plots from our main model showing logistic probability of a non-zero outcome for variables related to country-level surveillance and reporting. Light gray lines represent individual model iterations. Black center line is the median value and outer black lines are 95^th^ percentile ranges. Rug ticks show raw data values.

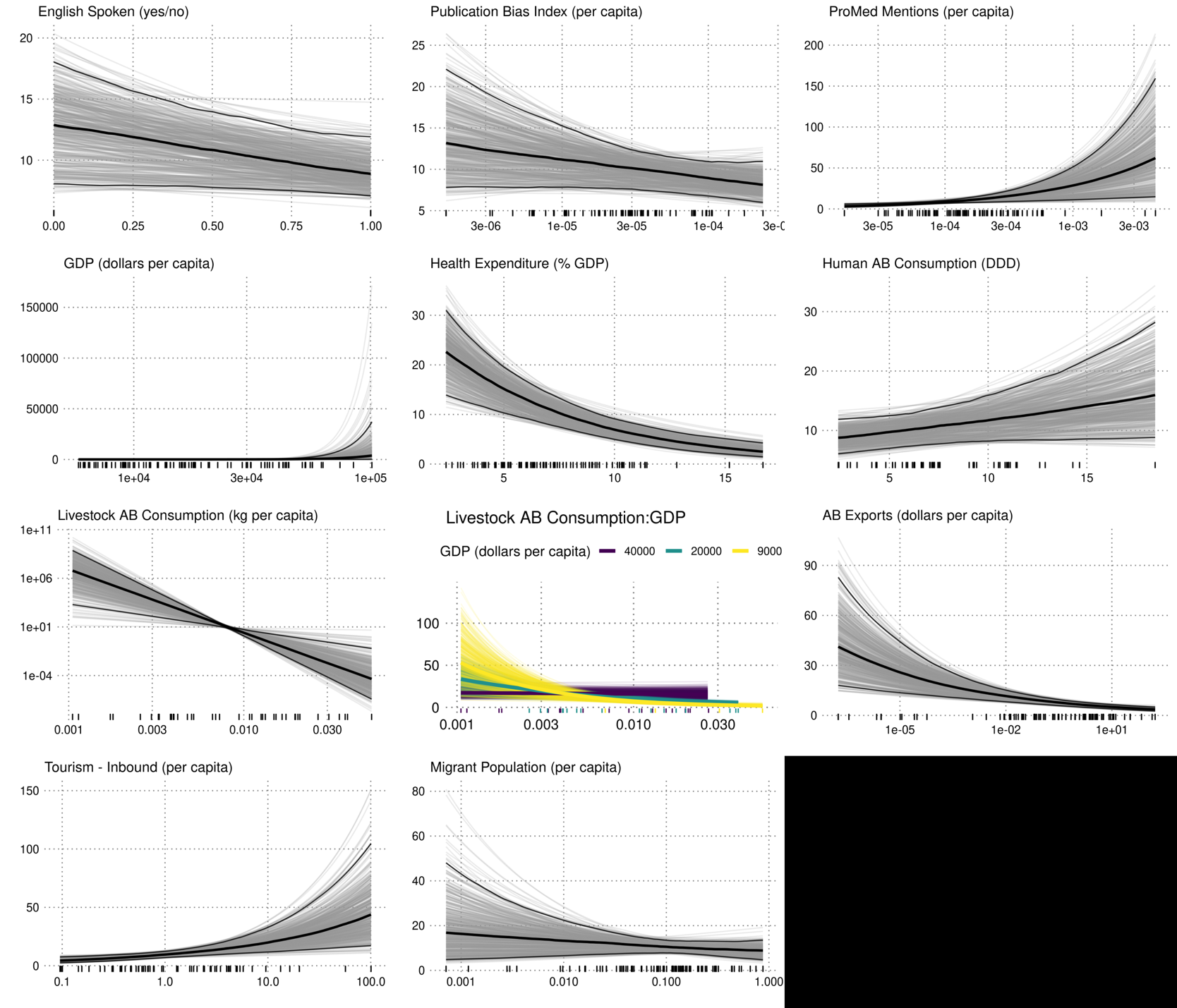

**Figure S1-b.** Partial effect plot showing predicted Poisson counts of AMR emergence events, conditional on observed reporting, for all variables in our main model. Light gray lines represent individual model iterations. Black center line is the median value and outer black lines are 95^th^ percentile ranges. Rug ticks show raw data values.

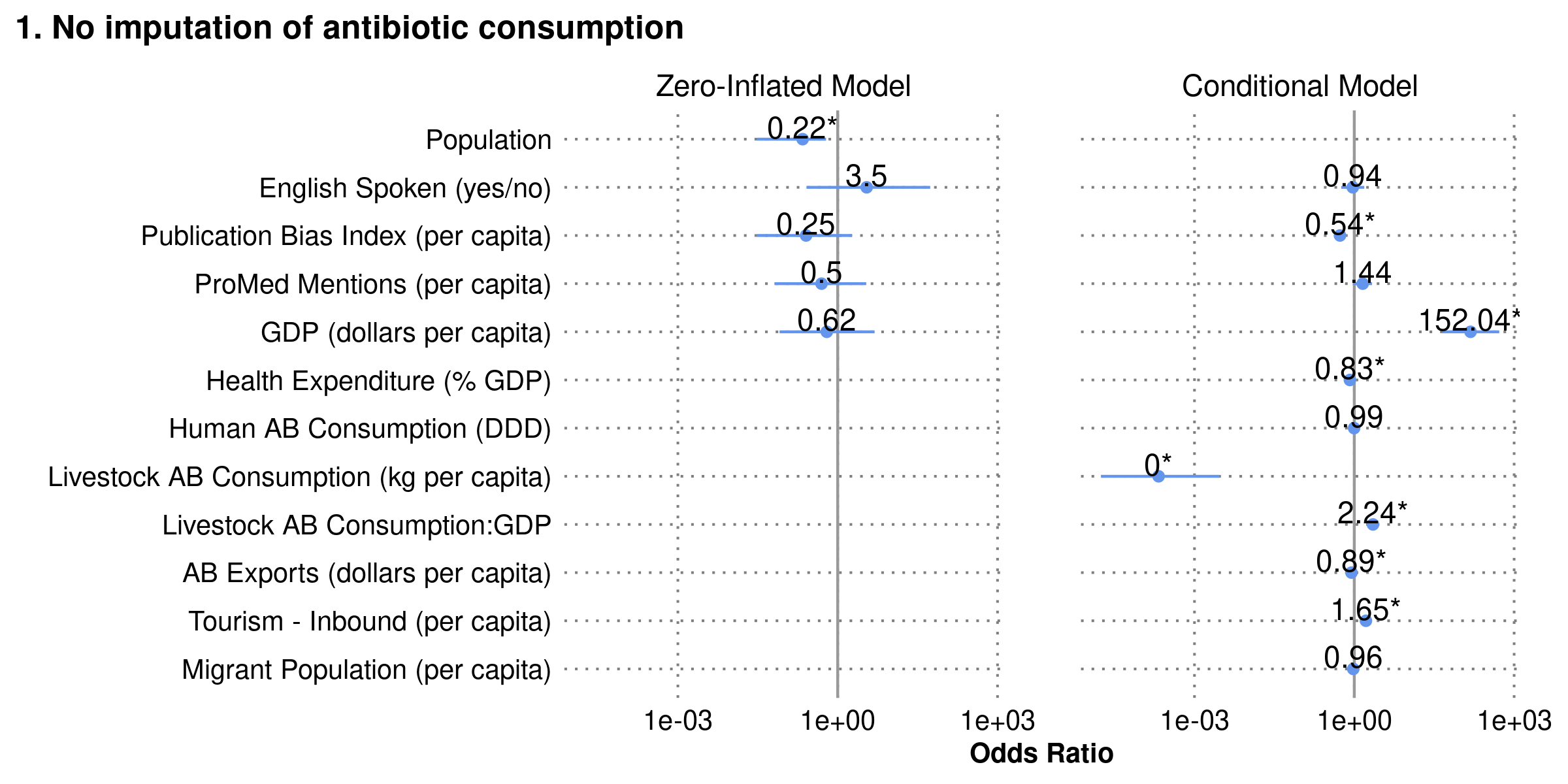

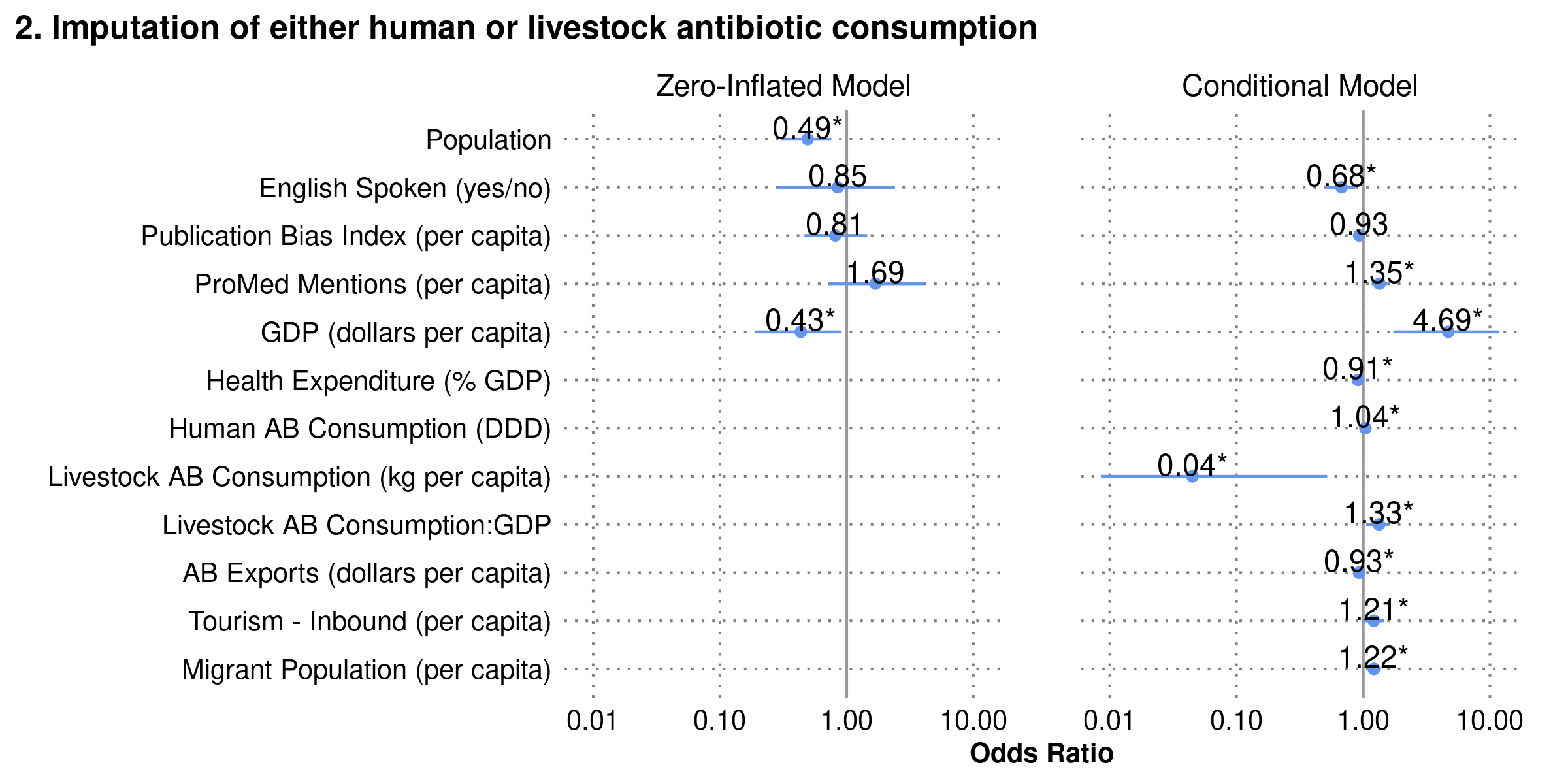

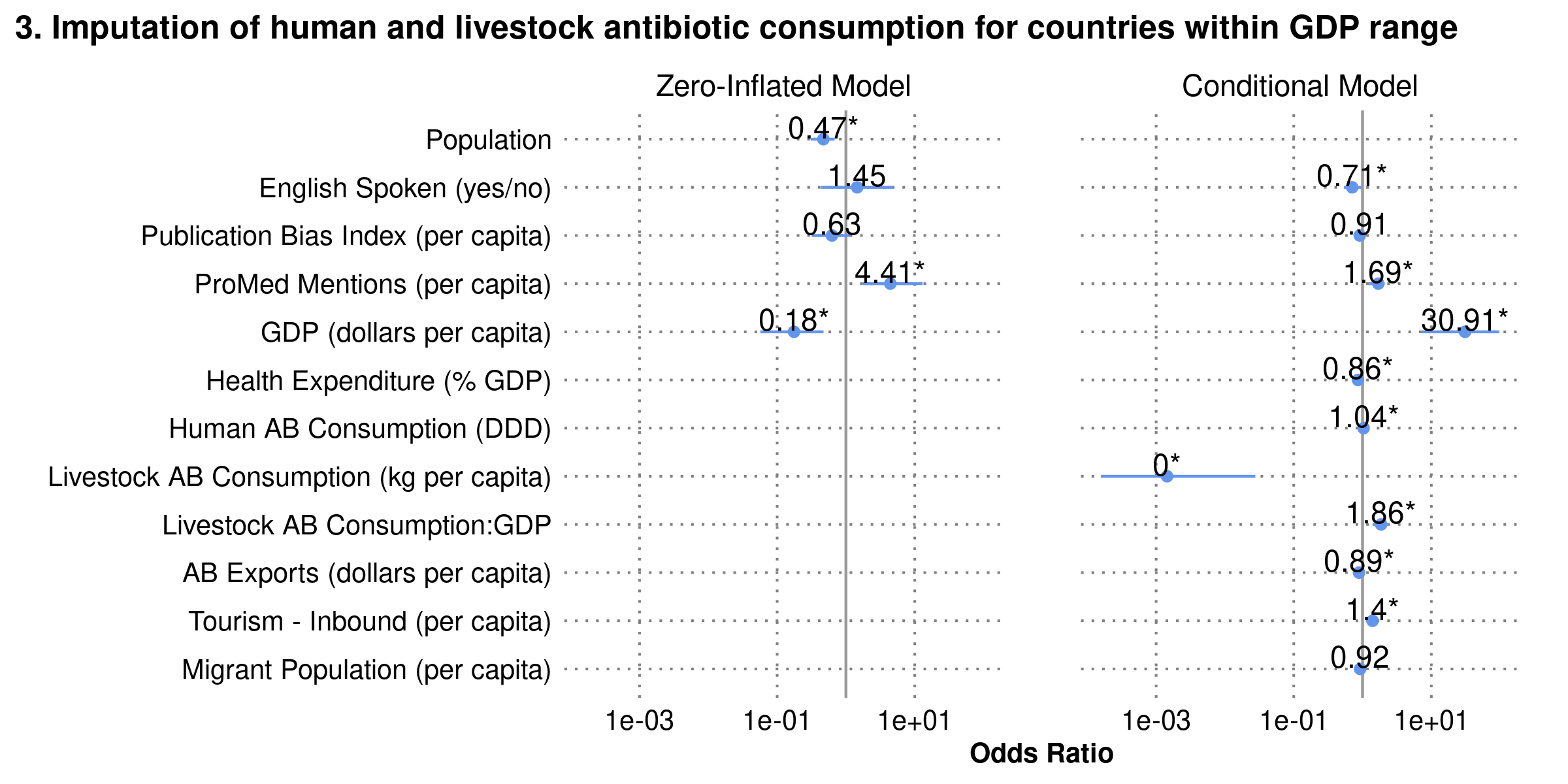

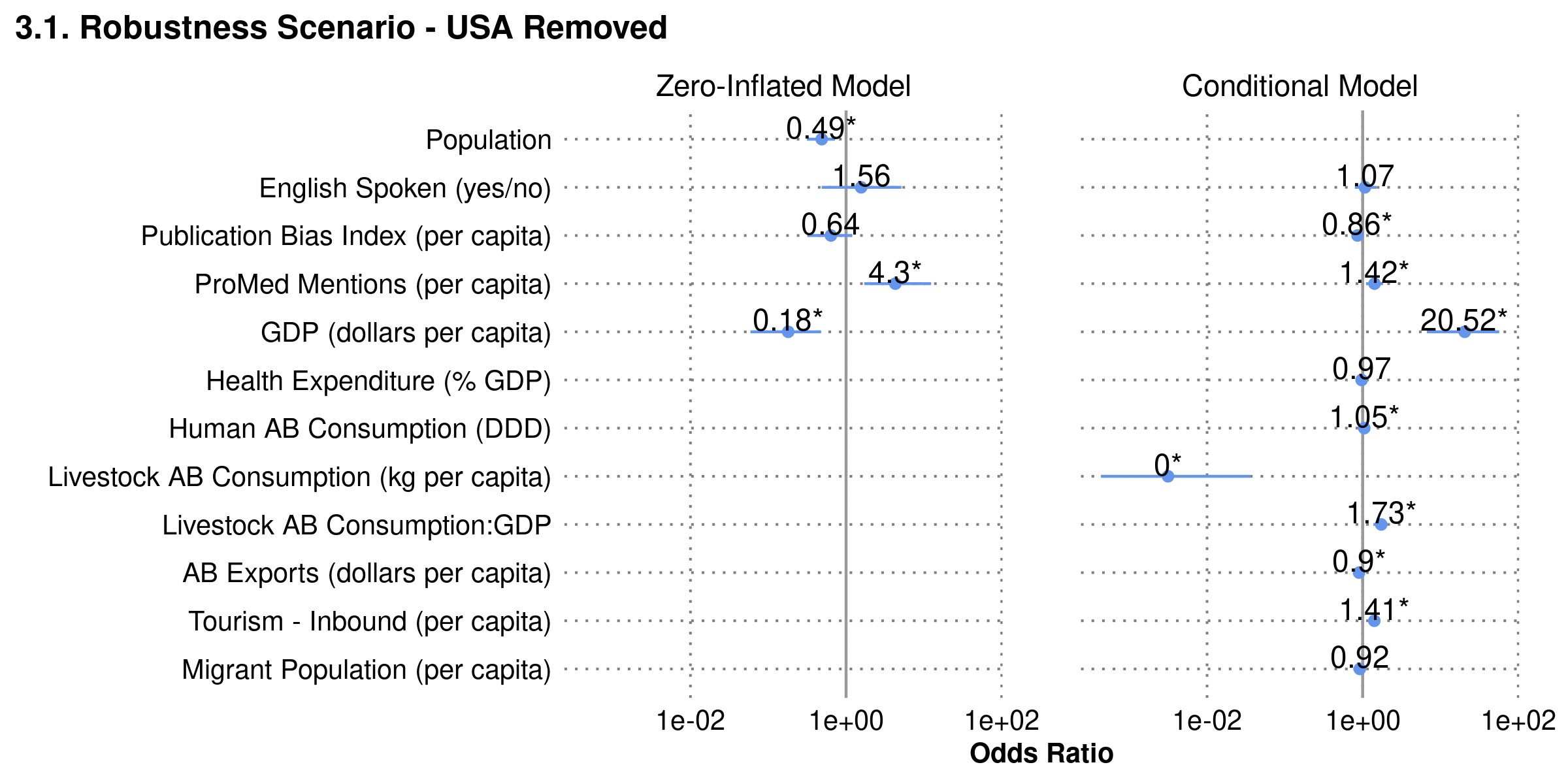

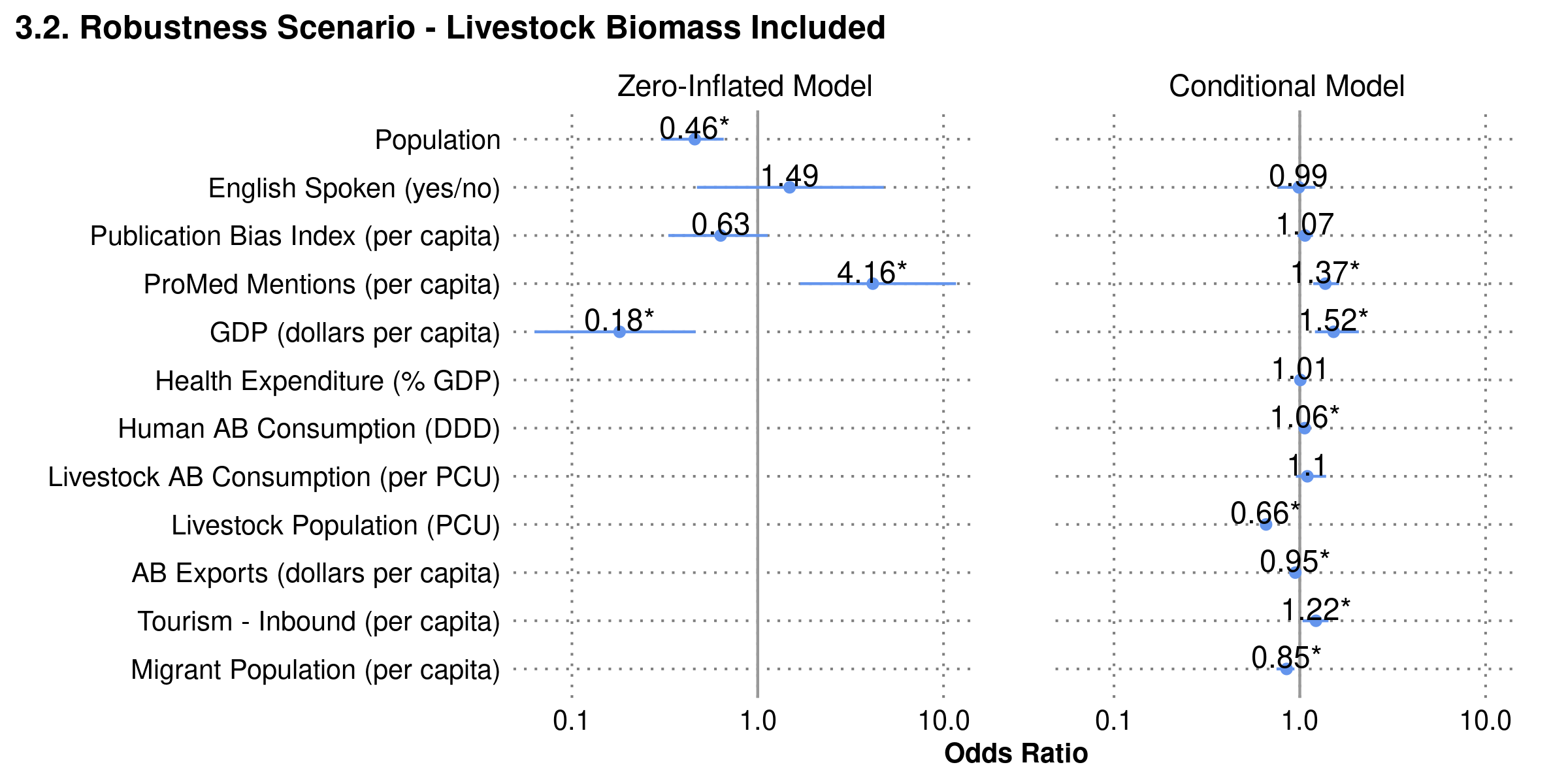

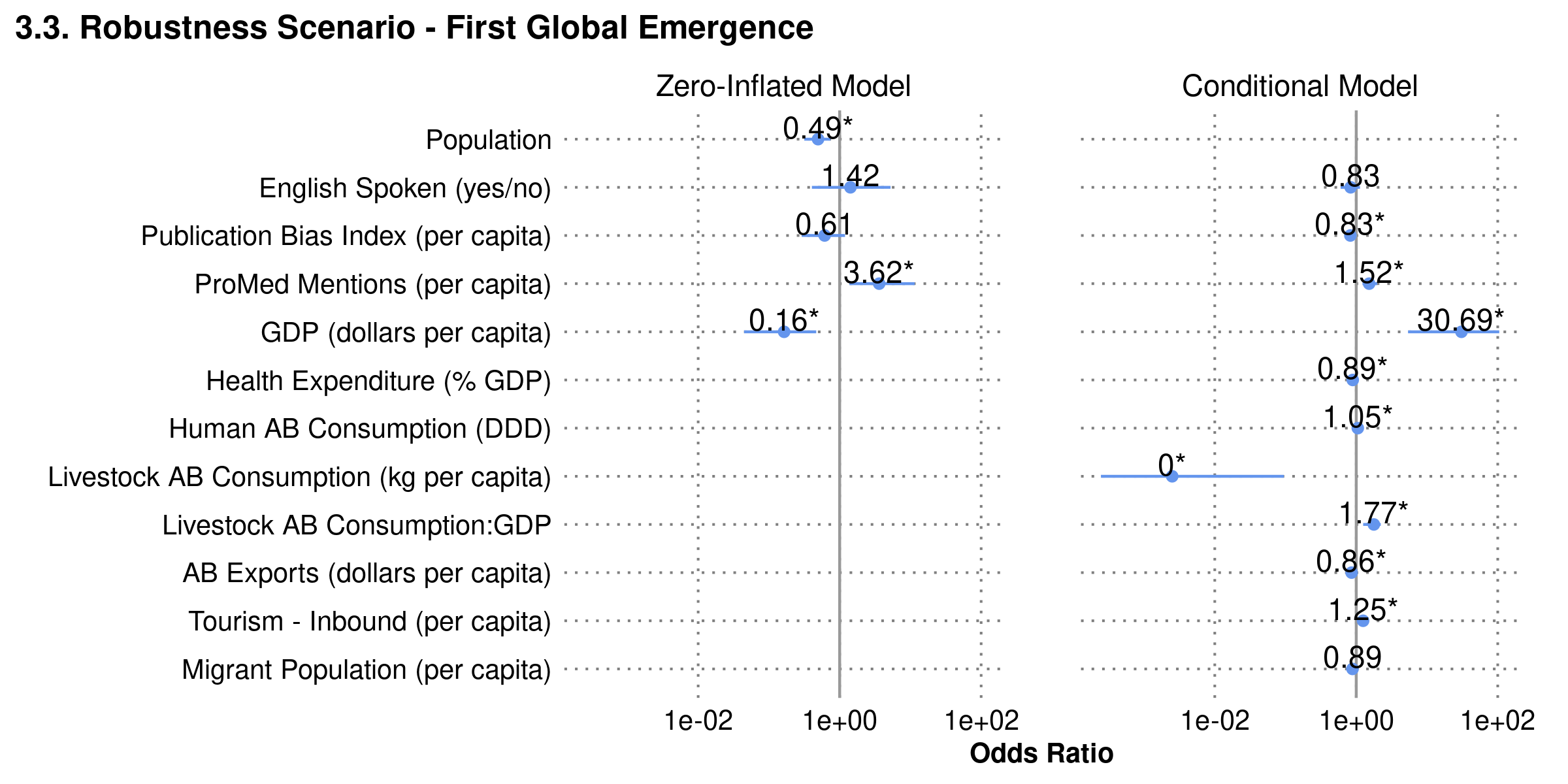

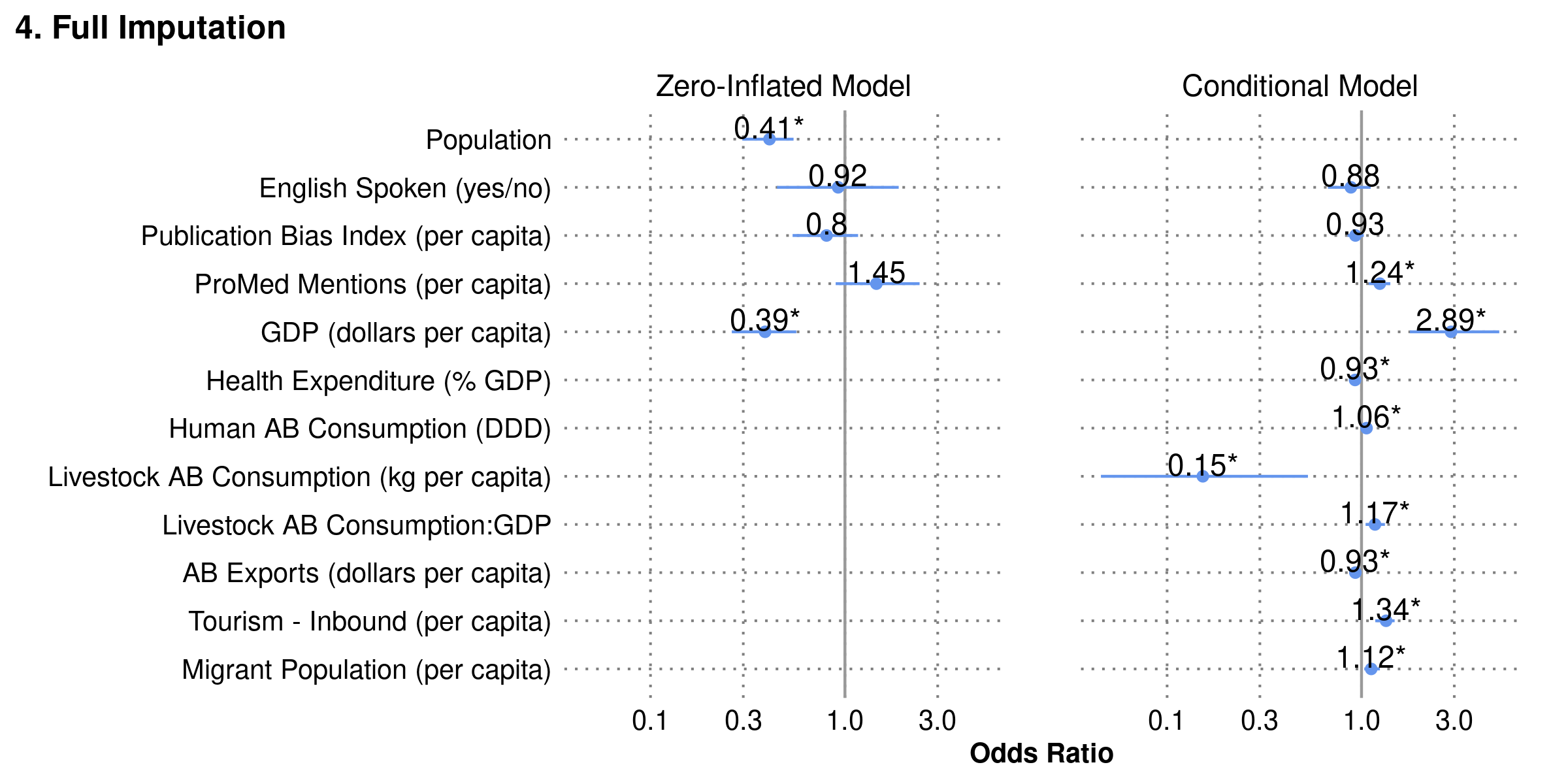

**Figure S2.** Odds ratios of features in alternative formulations of the model to test missing data imputation scenarios and model robustness. Model 3 (Imputation of human and livestock antibiotic consumption for countries within GDP range) is the main model, and is shown as **Figure 1** in the main text.

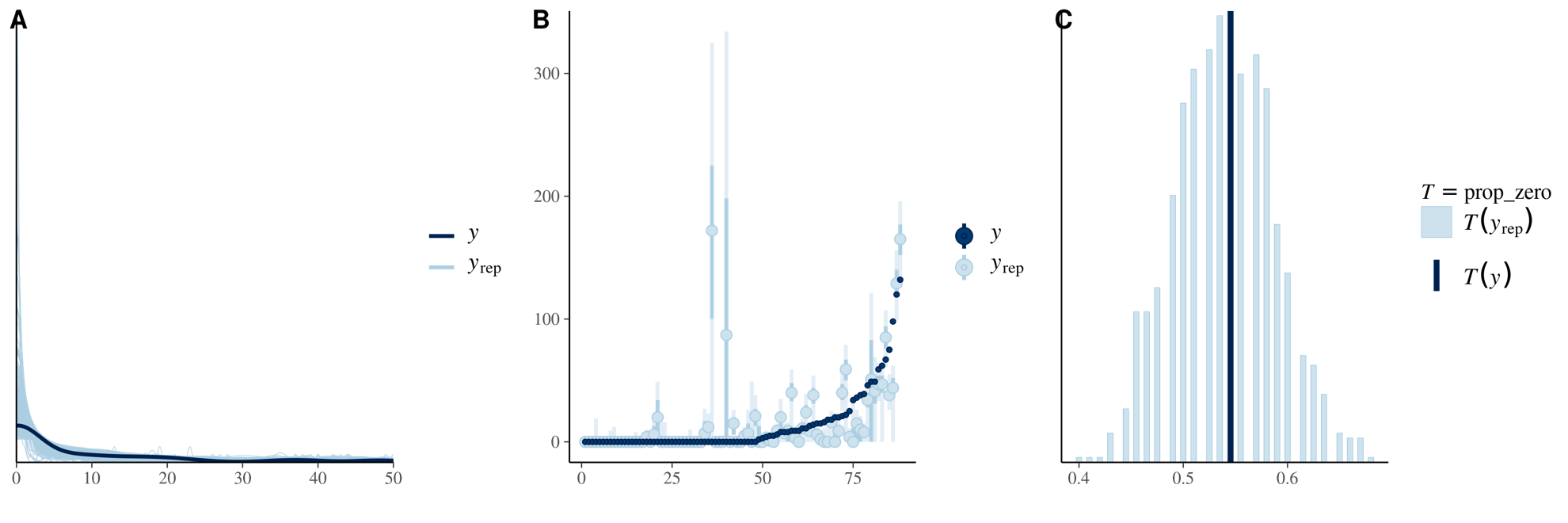

**Figure S3.** Model diagnostics for our main model. (A) density plot comparing empirical distribution of the data *y* to the distributions of simulated data *yrep* from the posterior predictive distribution. X-axis truncated at 50. (B) interval plot comparing empirical data points *y* to posterior predicted values for each point *yrep*. Dark green line represents 50% probability and faded line represents 90% probability. 36% non-zero empirical values are in 50% probability range and 58% non-zero empirical values in 90% probability range. (C) comparison of proportion of zeros computed from the empirical data *T(y)* to the distribution of proportion of zeros *T(yrep)* in the posterior datasets.
